# Can the Chest X-ray Brixia Score predict COVID-19 disease severity and mortality in resource-limited settings? A retrospective cross-sectional study at a tertiary hospital in Northwestern, Tanzania

**DOI:** 10.1101/2025.05.01.25326785

**Authors:** Dominic Mbwilo, Ziad Byekwaso, Ernest Elisenguo, Fredy Hyera, Evarist Msaki, Ally Munir Akrabi, Jeremiah Seni, Bahati Wajanga, Patrick Ngoya

## Abstract

**Background:** COVID-19 primarily affects the lung, thus the chest x-ray (CXR) is the first line imaging modality of evaluating the disease. CXR scoring systems for quantifying the severity and progression of lung abnormalities in COVID-19 pneumonia have been introduced. However, a chest X-ray severity scoring system has never been implemented in Tanzania to assist in predicting COVID-19 patient outcomes. Therefore, this study was designed to determine whether the CXR Brixia Score can predict outcomes among patients with COVID-19 pneumonia.

**Materials and Methods:** A retrospective cross-sectional hospital-based study conducted on COVID-19 pneumonia patients confirmed by nasopharyngeal swab RT-PCR assay at Bugando Medical Center (BMC) using data retrieved from the hospital database. Data retrieved included socio-demographics and clinical information. Baseline CXRs were reviewed and assigned a Brixia score by two experienced Radiologists. Disease severity, length of the hospital stay and in-hospital mortality were recorded as outcomes and correlated with the Brixia scores by logistic regression analysis after adjusting for potential cofounders. A p-value of <0.05 was significant.

**Results:** A total of 220 patients were enrolled with a mean age of 59 (±12.7) years. A severe Brixia score had a significant likelihood of severe form of disease (aOR=4.8, 95%CI=2.35 - 13.67, p=0.001) and death (aOR=5.39, 95%CI=1.65 - 24.73, p=0.041). A severe Brixia score did not predict a longer hospital stay (cOR=2.29, 95%CI=0.49 - 9.72, p=0.293).

**Conclusion:** CXR Brixia Score can predict disease severity and in-hospital mortality particularly in resource-limited settings. It may assist in timely identifying patients in need of aggressive management, and thereby avert adverse outcomes including mortality.

## Introduction

Globally, COVID-19 has been reported in many countries with a total 777,664,564 confirmed cases, 7,090,480 deaths by March 2025 with at least 67% of world population vaccinated [1].COVID-19 pneumonia may present various degrees of severity and respiratory impairment, ranging from mild cases with flu-like symptoms to severe forms with acute respiratory distress syndrome (ARDS), requiring intensive care support. Despite a potential multi-systemic involvement, the lung is by far the organ most commonly affected by SARS-CoV-2 infection.

Imaging is important in evaluating the extent, complications and follow up of COVID-19 pneumonia. The chest x-ray (CXR) has a relative high speed of acquisition, low cost, portability, and accessibility, especially in low-resource settings [2,3]. To improve the risk stratification, CXR scoring systems for quantifying the severity and progression of lung abnormalities in COVID-19 pneumonia were introduced [4]. The Brixia Score has the ability to provide relevant information for clinicians as well as identify the highest risk patients and those who require specific treatment strategies [5,6].The Brixia Score correlates positively with biomarkers of inflammation and organ injury known to be associated with severe COVID-19 [7].

In Tanzania, COVID-19 burden is alarmingly high especially among patients with severe and critical disease who have been documented to have a 31.8% to 34% mortality rate [8,9]. However, limited information still exists on the diagnostic utility of the baseline CXRs in identifying, exploring and categorizing COVID-19 patients so as to guide proper management, prevent deterioration, and predict long term complications and mortality in resource-limited settings. Therefore, this study was designed to determine whether the CXR Brixia Score can predict outcomes among patients with COVID-19 pneumonia.

## Materials and Methods

### Study design and setting

This was a retrospective cross-sectional hospital-based conducted on COVID-19 pneumonia patients data. COVID-19 diagnosis confirmed by real time-Polymerase Chain Reaction (rt-PCR) assay at BMC, Mwanza Tanzania using data accessed from the BMC Emergency Preparedness and Response Team database between 19 June to 30 September 2023 for analysis.

### Study population

All patients from the third and fourth waves of the COVID 19 pandemic diagnosed between 1^st^ September 2021 and 31^st^ May 2022. Patients with incomplete data were excluded.

### Study variables

Data retrieved included age, sex, underlying comorbidities, vital signs namely percentage oxygen saturation in ambient room air, respiratory rate (breaths per minute), pulse rate (beats per minute), systolic blood pressure (SBP) and diastolic blood pressure (DBP) in millimeters of mercury (mmHg) were recorded as exposures.

Disease severity, length of the hospital stay and in-hospital mortality were recorded as outcomes and correlated with the Brixia scores. Disease severity was categorized as; mild (no features suggestive of pneumonia by imaging diagnosis), moderate (with fever, respiratory tract symptoms and features suggestive of pneumonia by imaging diagnosis, SPO2 ≥ 94% on room air), severe (respiratory rate of 30 cycles /min, oxygen saturation ≤ 93% at rest, or lung infiltrates > 50%), and critical (patients with either respiratory failure requiring mechanical ventilation, in shock or requires ICU care for organ failure management). These were further re-categorized as; non-severe form of disease if mild and moderate severe disease or severe form of disease if severe and critical disease. Length of hospital stay was categorized as; short stay (<14 days) and long stay ≥ 14 days.

### Data collection and procedures

Nasopharyngeal swabs were collected at time of admission at EMD by qualified and trained clinical laboratory personnel and sent to BMC laboratory for processing using viral transport media. Abbott m2000 rt-PCR assay (Abbott Laboratories, USA) was used to detect RNA in respiratory specimens collected from patients who are suspected of COVID-19. All COVID-19 patients were subjected to routine CXRs using a XR6000 digital radiography system (GE Healthcare, USA). Baseline CXRs were reviewed and assigned a Brixia score by two experienced Radiologists. The Brixia chest X-ray severity scoring system was used to grade CXR abnormalities due to COVID-19 pneumonia into sum scores of 0 to 6 (mild), 7 to 12 (moderate) and ≥12 (severe) lung involvement [5,6].

### Data analysis

Data entry was done using Microsoft Excel and cleaned then exported to STATA version 15 for analysis. Continuous variables were summarized as mean with standard deviation (SD) or median with interquartile range (IQR) depending on the distribution. Categorical variables were summarized using frequency and proportions. COVID-19 pneumonia exposures and outcomes were analyzed and the strength of the association was assessed using logistic regression analysis before and after adjusting for potential cofounders. A p-value of <0.05 was significant.

### Ethical considerations

Ethical clearance for this retrospective study including waiver of informed consent was sought from and approved by the CUHAS/BMC Research and Ethical Review Committee (certificate number: CREC/680/2023) as well as from BMC. Confidentiality of data was strictly maintained by anonymization of data with restricted access to information that could identify individual participants during or after data collection.

## Results

Overall, 298 patients were confirmed with COVID-19 pneumonia during the study period. Seventy-eight patients were excluded due to incomplete clinical data (26.2%). Thus, 220 patients were enrolled in the study with an age range from 22 to 98 years and a mean age of 59 (±12.7) years. Around 121 patients were males (M:F = 1.2:1). About 114 (55%) had comorbidities, with the majority being hypertensive (40%). The median (IQR) oxygen saturation in ambient room air at the admission was 85% (76-91), the lowest and highest recorded values being 34% and 97%, respectively. The median (IQR) of hospital stay of enrolled patients was 9 (6-13), the lowest stay being 2 days and the longest stay being 71 days. The distribution of lung involvement was assigned a Brixia score at the time of admission, whereby mild lung involvement was seen in 12 patients (5.5%), as shown by an example of a CXR with a Brixia score of 4 (**Figure 1**). Moderate lung involvement was seen in 57 patients (25.9%), as shown by an example of a CXR with a Brixia score of 8 (**Figure 2**). Severe lung involvement was seen in 151 patients (68.6%), as shown by an example of a CXR with a Brixia score of 14 (**Figure 3**). Most of the patients (70.6%) had a severe disease with 32 patients (14%) who were in critical condition required ICU admission (**Table 1**). The severe forms of COVID-19 disease among patients were significantly predicted by severe Brixia score (aOR=4.8, 95%CI=2.35 - 13.67, p=0.001) (**Table 2**). Similarly, death was also significantly predicted by severe Brixia score (aOR=5.39, 95%CI=1.65 - 24.73, p=0.041) (**Table 3**). A severe Brixia score did not predict a longer hospital stay (cOR=2.29, 95%CI=0.49 - 9.72, p=0.293) (**Table 4**).

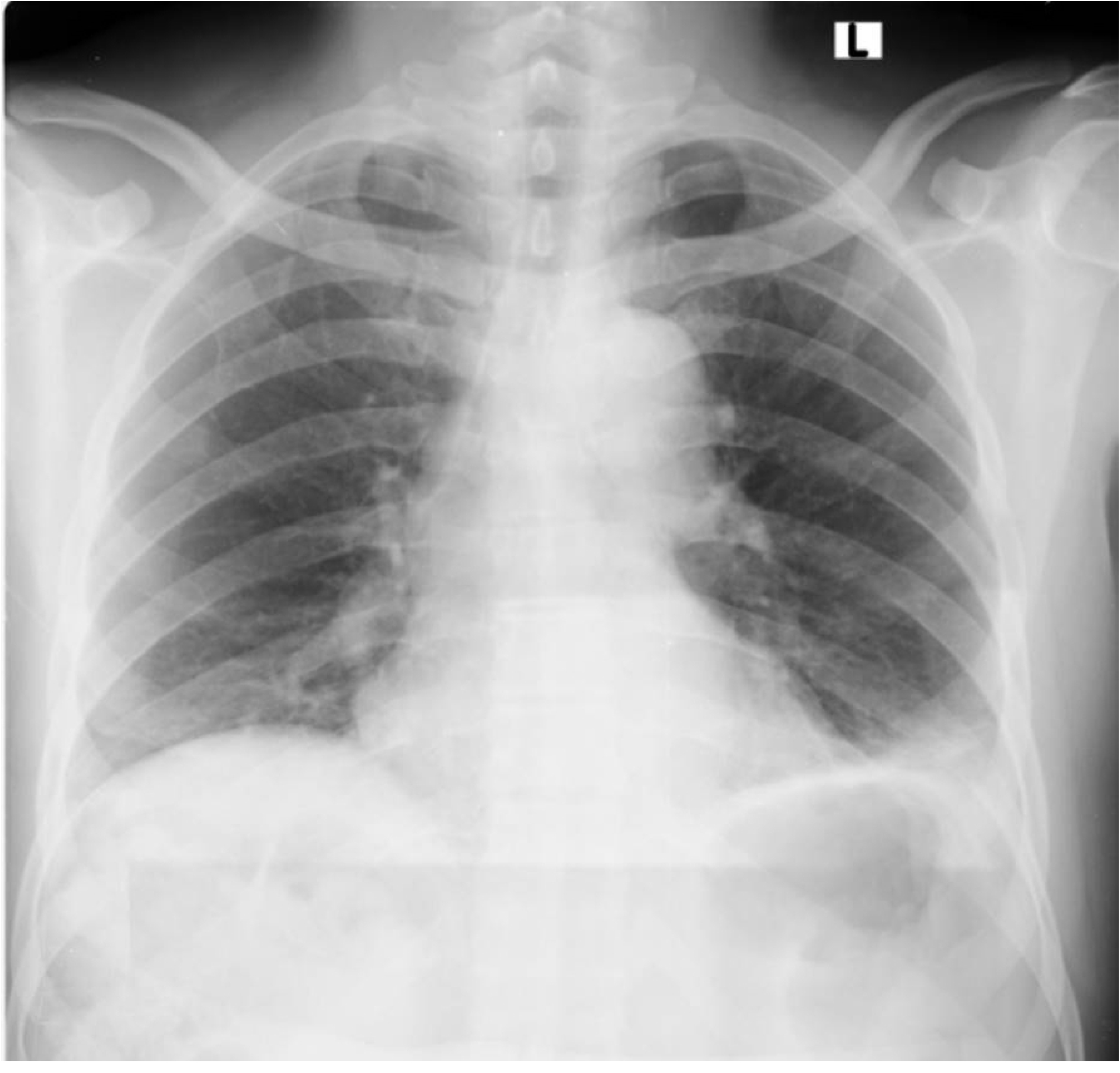

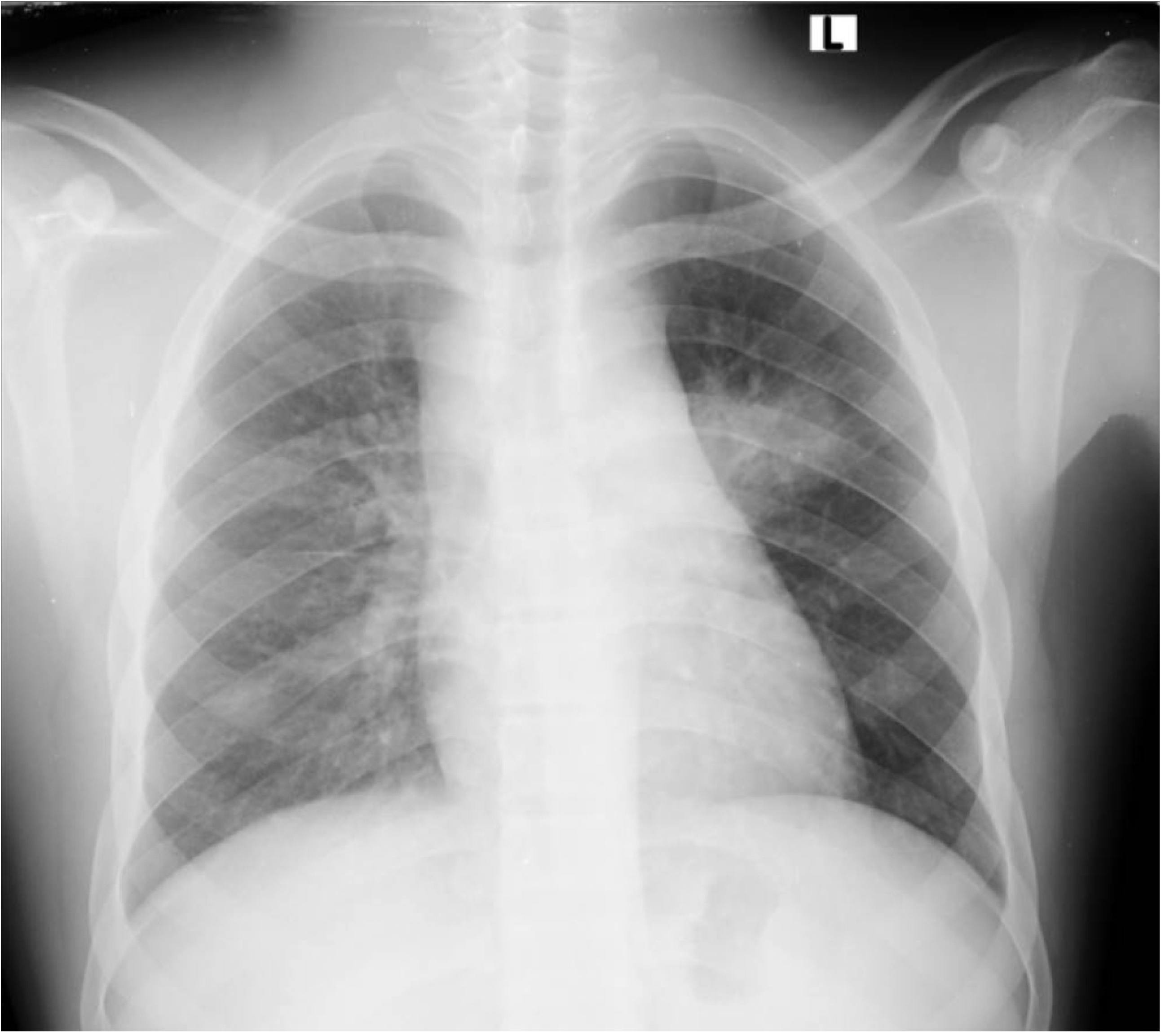

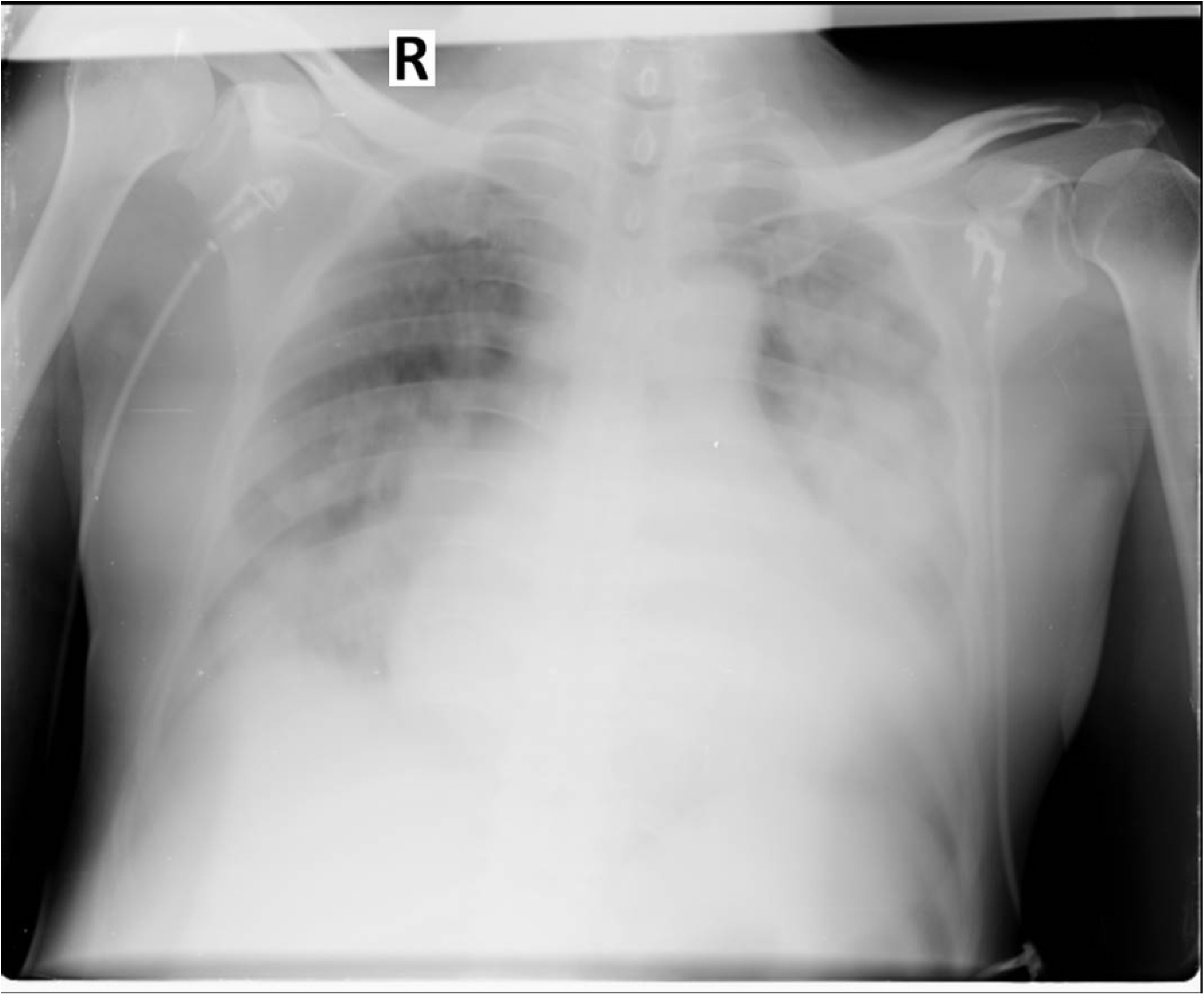

**Table 1.** Social demographic and clinical characteristics of study participants (n=220).

| Variable | n (%) |
| --- | --- |
| Age mean (SD) in years | 59 (±12.7)<sup>#</sup> |
| Age (<60 years) | 112 (50.9) |
| Sex (Males) | 121 (55.0) |
| Comorbidities (Yes)* | 121 (55.0) |
| Hypertension | 48 (22.0) |
| Diabetes Mellitus | 23 (10.5) |
| Chronic Kidney Disease | 11 (5) |
| Cough (Yes) | 190 (86.4) |
| Difficulty in Breathing (Yes) | 166 (72.7) |
| Oxygen saturation median (IQR) | 85 (76-91)<sup>#</sup> |
| Temperature median (IQR) | 37.2 (36.7-37.9)<sup>#</sup> |
| Respiratory rate median (IQR) | 28 (24-32)<sup>#</sup> |
| Pulse rate median (IQR) | 92 (81-102)<sup>#</sup> |
| Systolic BP median (IQR) | 126 (111-145) |
| Diastolic BP median (IQR) | 76 (66-87)<sup>#</sup> |
| Mild lung involvement (0 to 6)* | 12 (5.5) |
| Moderate lung involvement (7 to 12)* | 57 (25.9) |
| Severe lung involvement (≥12)* | 151 (68.6) |
| Mild to moderate disease | 66 (30.0) |
| Severe to critical disease* | 154 (70.0) |
| Length of the hospital stay median (IQR) | 9 (6-13)<sup>#</sup> |
| In-hospital mortality | 68 (30.9) |

<sup>#</sup>Non-categorical variables, \*Brixia Score; \*\*Intensive Care Unit (ICU) admission (14.2%).

**Table 2.** Association between baseline Brixia Score and disease severity (n=220).

| Brixia Score | Disease severity |  | Univariate |  |  | Multivariate |  |  |
| --- | --- | --- | --- | --- | --- | --- | --- | --- |
|  | Mild-Moderate | Severe-Critical | cOR | 95% CI | p | aOR | 95% CI | p |
| Mild | 10 (15.6) | 2 (1.3) | 1 |  |  |  |  |  |
| Moderate | 36 (54.1) | 21 (13.6) | 2.65 | 0.83-8.41 | 0.098 | 2.48 | 0.66- 9.34 | 0.177 |
| Severe | 20 (30.3) | 131 (85.1) | 6.27 | 3.23 - 10.26 | **<0.001** | 4.8 | 2.35 - 13.67 | **0.001** |

cOR = crude Odds Ratio, CI=Confidence Interval, p=p-value, aOR = adjusted Odds Ratio

**Table 3.** Association between baseline Brixia Score and in-hospital mortality (n=220).

| Brixia Score | In-hospital mortality |  | Univariate |  |  | Multivariate |  |  |
| --- | --- | --- | --- | --- | --- | --- | --- | --- |
|  | Survived | Deceased | cOR | 95% CI | p | aOR | 95% CI | p |
| Mild | 11(7.4) | 1(1.4) | 1 |  |  |  |  |  |
| Moderate | 49(32.2) | 8(11.7) | 3.1 | 0.64 - 11.08 | 0.13 | 4.24 | 0.50-19.84 | 0.185 |
| Severe | 92(60.54) | 59(86.9) | 4.21 | 1.04-13.61 | **0.02** | 5.39 | 1.65-24.73 | **0.041** |

cOR = crude Odds Ratio, CI=Confidence Interval, p=p-value, aOR = adjusted Odds Ratio

**Table 4.** Association between baseline Brixia Score and the length of hospital stay (n=220).

| Brixia Score | Length of hospital stay |  | Univariate |  |  |
| --- | --- | --- | --- | --- | --- |
|  | <14 days<br>n (%) | ≥ 14 days<br>n(%) | cOR | 95% CI | p |
| Mild | 10 (6.3) | 2(3.1) | 1 |  |  |
| Moderate | 39(24.6) | 18(29.8) | 1.05 | 0.21-5.27 | 0.945 |
| Severe | 109(68.1) | 42(67.1) | 2.29 | 0.49-9.72 | 0.293 |

cOR = crude Odds Ratio, CI=Confidence Interval, p=p-value, aOR = adjusted Odds Ratio

## Discussion

This study found remarkably high adverse outcomes among patients with COVID-19 pneumonia, which included severe disease (70.0%), mortality (30.9%) and prolonged length of hospital stay (28.2%). These adverse outcomes are similar to other studies done in Tanzania, which altogether calls for prompt risk stratification of patients upon admission, expediting diagnostics and fostering aggressive management [8,9]. On multivariate analysis, there was a significant likelihood of severe form of COVID-19 pneumonia if a patient had a severe Brixia score. This implied that a higher Brixia score on admission increased the risk of developing severe form of COVID-19. This is in line with a study done in Finland that included of 37 hospitalized COVID-19 patients with 290 CXRs and another retrospective study done in Italy involving 524 CXRs of COVID-19 patients which showed that a Brixia score at time of admission correlated with severity of disease and outcome [7,10]. Severe lung involvement in patients with severe form of disease is attributed by pathogenesis of COVID-19, as SARS-CoV2 favor type II cells over type I cells when infecting the lung thus peripheral and sub pleural alveolar units within the lung are primary affected and lead to typical features of SARS-CoV2 on imaging. SARS-CoV2 spreads in type II cells, and releases a considerable amount of viral particles before the cells triggering an inflammatory response ear marked by cytokine storms and ultimately the cells undergo apoptosis [11]. Diffuse alveolar destruction with fibrin-rich hyaline membranes and some multinucleated large cells is the pathologic outcome of SARS-CoV2 in COVID-19 pneumonia account for the typical features seen in the CXRs [12]. Therefore, capitalizing in these hallmarks to guide COVID-19 patients’ risk stratification is reiterated in our findings, and a pressing need to integrate them in country-specific radiological screening and diagnostics algorithms.

Death was significantly likely on multivariate analysis if patient had a severe Brixia score compared to the survival group. This is similar to a retrospective study done in Italy on 302 COVID-19 pneumonia patients to predict the risk of in-hospital mortality which demonstrated that patients with a high Brixia score and at least one other predictive factor had the highest risk of in-hospital death [13]. Another retrospectively study reviewed 125 baseline CXRs of adults with COVID-19 confirmed patients admitted at a private tertiary hospital in Philippines suggested that Brixia scores obtained from baseline CXR have a significant association with in-hospital mortality [14].

In this study, we observed no significant likelihood of a longer hospital stay for patients with COVID-19 pneumonia if patient had a severe Brixia score. Similar findings were seen in a case-control study from University Islam Bandung, Indonesia involving 653 COVID-19 confirmed patients showed that CXR Brixia score can predict mortality, but it cannot predict the length of stay of hospitalized COVID-19 confirmed patients [15].

Our findings highlight a need to review country-specific screening algorithms for COVID-19 patients on admission. It will be prudent to include diagnostic parameters such as the CXR in addition to the existing clinical and laboratory parameters to foster reliable disease prediction and thereby favoring better patients’ management outcomes. It is worth noting that in this study only a single baseline Brixia Score was recorded rather than serial follow up Brixia Scores which may be of interest in future studies.

## Conclusions

CXR Brixia Score can predict disease severity and in-hospital mortality particularly in resource-limited settings. It may assist in timely identifying patients in need of aggressive management, and thereby avert adverse outcomes including mortality.

## Data Availability

Data are available from the BMC Emergency Preparedness and Response Team database (contact via email) for researchers who meet the criteria for access to confidential data.

## Acknowledgement

The authors would like to thank the BMC Emergency Preparedness and Response Team for providing the technical support to this study.

## Authors’ contributions

Conceived, designed and executed the study (DM, ZB, EE, AMR, JS, BW and PN). Supervised the study, and coordinating the COVID-19 database at Bugando (ZB, EE, AMA, JS, BW and PN). Data and sample collections (DM, ZB, EE, FH and EM). Patients’ management (DM, ZB, EE, AMR, JS, BW and PN). Data analysis, literature search and interpretation of data (DM, ZB, EE, FH, EM, AMR, JS, BW and PN). Preparation of the first draft of the manuscript (DM and PN). Critical review of the manuscript (DM, ZB, EE, FH, EM, AMR, JS, BW and PN). All authors read and approved the final manuscript.

## Availability of data and materials

Data is available from the authors upon request.

